# Identification of key genes involved in neuroendocrine regulation in pulpitis: bioinformatics and experimental analysis

**DOI:** 10.64898/2026.04.18.26351158

**Authors:** Huimin Jin, Yawei Wang, Aoteng Sun, Yun Liu, Ting Guo

## Abstract

**Background:** There is a close correlation between neuroendocrine regulation and pulpitis progression. This study aims to identify key neuroendocrine regulation-related genes in pulpitis, providing insights for its treatment.

**Methods:** GSE77459 and GSE92681 datasets were used to validate experimental findings. Key neuroendocrine regulation-related genes were identified via Cytoscape plugin cytoHubba and expression validation. Gene set enrichment analysis, RNA-binding protein regulatory networks, post-translational modifications, molecular regulatory networks, and drug prediction were performed. Key gene expression was experimentally verified in clinical samples.

**Results:** Top 10 genes were obtained via cytoHubba; 4 (IL6R, OSM, IL1RN, CCL4) with significant differences between pulpitis and control samples and consistent trends in both datasets were identified as key genes. Gene set enrichment analysis showed key genes participate in pathways like cytokine-cytokine receptor interaction. Related RNA-binding proteins were ELAVL1 and HNRNPA1, with phosphorylation as the main post-translational modification. Core regulatory microRNAs included miR-519, miR-765, miR-23, and regulatory factors included FOXC1, PRRX2. Targeted drugs (e.g., sarilumab, haloperidol decanoate, cyclosporine) were predicted, and clinical sample verification confirmed consistent expression trends.

**Conclusion:** 4 key neuroendocrine regulation-related genes were identified, which may have clinical significance for the diagnosis and treatment of pulpitis.

## 1. Introduction

Pulpitis is an inflammatory state of the dental pulp, mainly caused by bacterial invasion resulting from untreated caries, traumatic injuries, or iatrogenic damage during dental procedures.[1, 2] Clinically, it is characterized by spontaneous or stimulus-induced pain, thermal sensitivity, and in some cases, localized swelling. If left untreated, it may progress to pulp necrosis, periapical pathology, or more severe complications.[3–5] Reversible pulpitis is typically managed through caries removal and protective treatments such as indirect pulp capping and stepwise excavation, while irreversible pulpitis requires more invasive interventions, including root canal therapy or tooth extraction. Therefore, accurately assessing the severity of pulpitis is crucial for choosing an appropriate treatment plan. However, the clinical assessment mainly relies on the patient’s pain level, pulp sensitivity tests, and medical history,[6] which is not effective and precise enough, and may lead to incorrect judgment of the pulp status and inappropriate treatment measures. In light of these diagnostic challenges, the identification of key genes associated with pulpitis pathogenesis could potentially offer novel molecular biomarkers for more precise diagnosis and may contribute to the development of personalized treatment strategies.

Neuroendocrine regulation (NER) encompasses the complex bidirectional communication between the nervous and endocrine systems, involving both central structures (hypothalamus and pituitary gland) and peripheral tissues, to coordinate physiological responses through the release of neurotransmitters, neuropeptides, and hormones.[7–10] This process involves the interaction between the nervous system and the endocrine system, where the nervous system regulates the function of target cells by releasing neurotransmitters and hormones, maintaining the homeostasis of the internal environment.[7] NER plays pivotal roles in various physiological processes, including stress responses, immune system modulation, pain perception, and tissue repair mechanisms, all of which are particularly relevant in the context of inflammatory conditions.[11, 12]

The dental pulp, a highly innervated tissue with dense sensory nerve fibers and immune cells.[13] represents a potential hub for NER-mediated interactions during pulpitis. Sensory neurons in the pulp release neuropeptides like substance P and calcitonin gene-related peptide (CGRP), which directly amplify vascular dilation, immune cell infiltration, and pro-inflammatory cytokine release.[14] Conversely, inflammatory mediators during pulpitis may dysregulate neuroendocrine signals, sensitizing nociceptors and perpetuating pain.[15, 16] This bidirectional crosstalk suggests that NER imbalances might exacerbate pulpitis progression or impair reparative dentinogenesis. Understanding these complex neuroendocrine-immune interactions provides a foundation for developing novel therapeutic strategies that could potentially target these pathways to mitigate inflammation, alleviate pain, and preserve pulp vitality. Transcriptomic profiling of pulp tissues could unravel key genes and pathways linking NER to pulpitis pathogenesis, advancing personalized therapeutic approaches in endodontics.

This study is based on public datasets. Through differential expression analysis, differentially expressed genes related to neuroendocrine regulation were obtained. Key genes were acquired through protein interaction network construction and expression verification. Functional enrichment analysis of key genes, RBP network construction, post-translational modification analysis of proteins, molecular regulatory network construction, and drug prediction were conducted to explore the molecular mechanisms of key genes from various aspects. In conclusion, verification assays were performed to check if the expression of key genes matched the results of bioinformatic analyses, so as to supply novel support for the clinical diagnosis and therapy of pulpitis. The entire analysis procedure is shown in **Fig 1**.

**Fig 1.**
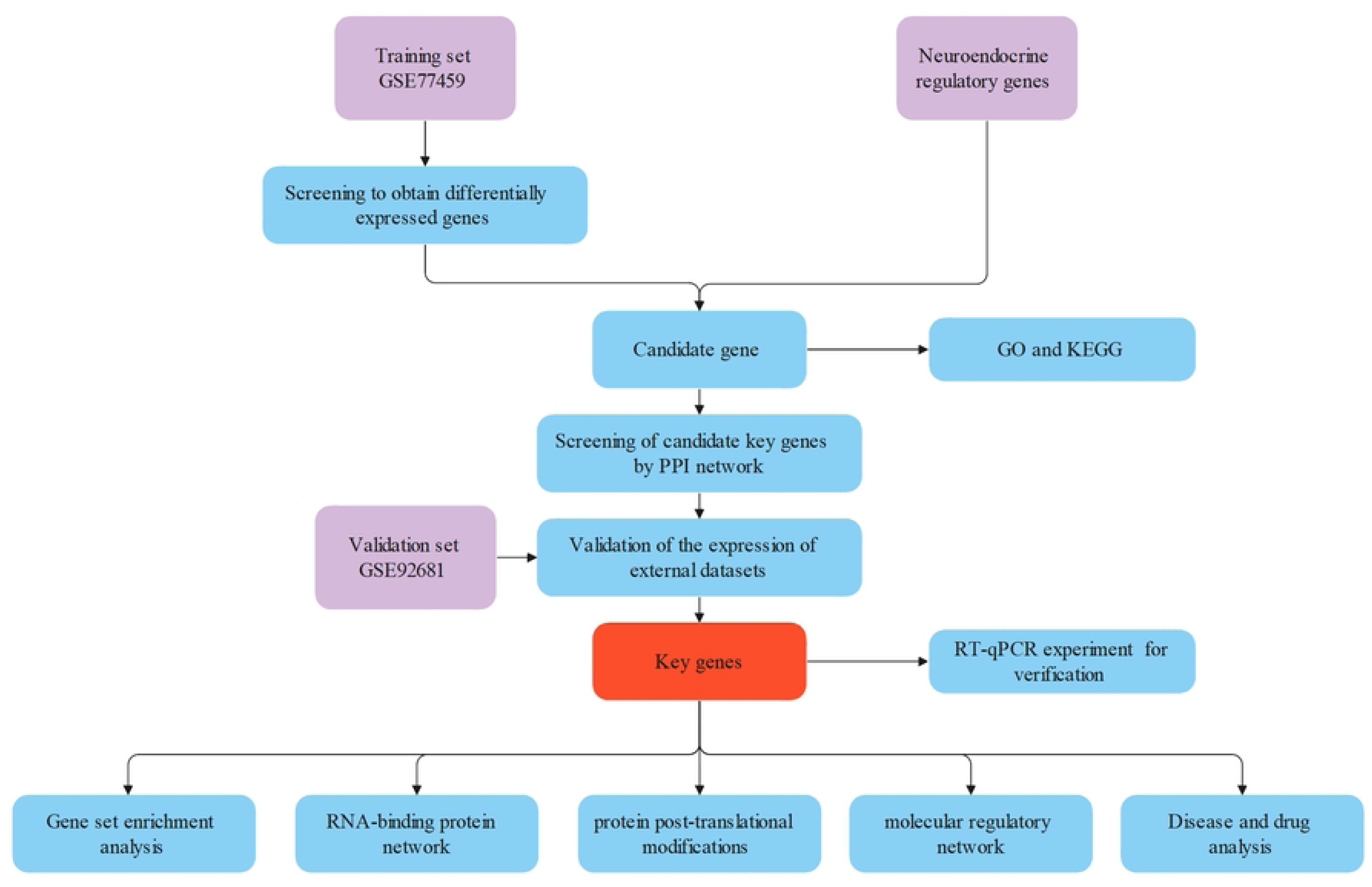
General workflow of this study.

## 2. Study materials and methodologies

### 2.1 Data retrieval and curation

Datasets associated with pulpitis were acquired from the Gene Expression Omnibus (GEO) repository (http://www.ncbi.nlm.nih.gov/geo/). Among them, the dataset GSE77459 (platform GPL17692) was composed of pulp tissue samples from 6 pulpitis patients and 6 controls, which was designated as a training set. The validation set GSE92681 (platform GPL16956) encompassed pulp tissue samples from 7 pulpitis patients and 5 controls. Furthermore, a total of 1,173 neuroendocrine regulation-related genes (NER-RGs) (**S1 Table**) were integrated from relevant literature.[17]

### 2.2 Assessment of differential gene expression patterns

To identify differentially expressed genes (DEGs) between pulpitis and control samples within the training set, the ‘limma’ package (v 3.54.0)[18] was employed for differential expression analysis, and DEGs were screened based on P < 0.05 and |log_2_ fold change (FC)| ≥ 1. Subsequently, the ‘ggplot2’ package (v 3.4.1)[19] was utilized to construct a volcano plot for visualizing the DEGs, and the ‘Heatmap’ package (v 1.0.12)[20] was used to generate a heatmap.

### 2.3 Identification and functional analysis of candidate genes

The DEGs and NER-RGs were subjected to intersection analysis using ‘ggvenn’ package (v 0.1.9)[21] was used to intersect the DEGs and NER-RGs, and and the overlapping genes were designated as candidate genes. Subsequently, Gene Ontology (GO) and Kyoto Encyclopedia of Genes and Genomes (KEGG) enrichment analyses were performed with the ‘clusterProfiler’ package (v 4.2.2)[22] (P < 0.05), showing the items with the most significant enrichment. In addition, the candidate genes were input into the STRING (confidence score > 0.4). A protein-protein interaction (PPI) network was visualised using the ‘Cytoscape’ software (v 3.8.2)[23]. Subsequently, the DMNC algorithm of the CytoHubba plugin in Cytoscape was employed to screen the top 10 genes in terms of score as candidate key genes.

### 2.4 Identification and validation of key genes

To observe whether the expression and trend of the candidate key genes were consistent in both the validation set and the training set, the expression levels of the candidate key genes in the pulpitis and control samples were compared by the Wilcoxon test in the training and validation sets (P < 0.05). Subsequently, the genes whose expression levels differed in the pulpitis and control samples and that had a consistent trend in the 2 datasets were selected and named as the key genes.

### 2.5 Gene set enrichment analysis (GSEA)

GSEA of key genes was performed, with the reference gene set ‘c2.cp.v7.2.symbols.gmt’ retrieved from the MSigDB. Firstly, based on all samples in the training set, the ‘psych’ package (v 2.1.6)[24] was utilized to conduct separate Spearman correlation analyses between each key gene and the remaining genes, yielding correlation coefficients. GSEA was then carried out on the sorted data (P valueCutoff = 0.05, P AdjustMethod = ‘BH’, minGSSize = 20, maxGSSize = 500) using the ‘clusterProfiler’ package (v 4.2.2)[22]. Finally, the ‘enrichplot’ package (v 1.18.3)[25] was utilized to visualize the significantly enriched pathways.

### 2.6 Analysis of RNA-binding protein (RBP) network and protein post-translational modifications

To understand the RBPs of key genes, the ENCORI database was used for prediction, and the Cytoscape software (v 3.8.2)[23] was utilized to construct the regulatory network of key gene-RBP.

To investigate the types of post-translational modifications and related sites of key genes at the protein level, PhosphoSitePlus (https://www.phosphosite.org/homeAction.action) was used for analysis.

### 2.7 Construction of molecular regulatory network

To probe the intricacy and multiplicity of the regulation process of key genes’ expression by constructing molecular regulatory networks, the miRNAs were forecasted using microcosm and diana-microt. The intersections of the predicted mRNA-miRNA pairs from the two databases were taken. Moreover, the lncRNAs upstream of the miRNAs were forecasted using the starBase. Besides, the Network Analyst database was employed to predict the transcription factors (TFs) of key genes, and the lncRNA-miRNA-mRNA as well as TF-mRNA networks were established with Cytoscape (v 3.8.2)[23].

### 2.8 Disease and drug analysis

To probe the potential drugs for treating pulpitis and study potential associations between key genes and drugs, the DGIdb (https://dgidb.org/) was employed to forecast drugs potentially connected with key genes. Besides, analysis of the relationship between key genes and diseases was performed with the Comparative Toxicogenomics Database (CTD, https://ctdbase.org/). Finally, visualization of the drug-key gene-disease network was accomplished via ‘Cytoscape’ software (v 3.8.2)[23].

### 2.9 Clinical sample validation

In this study, eight human dental pulp tissue samples were obtained from the Nanjing Stomatological Hospital, Affiliated Hospital of Medical School, Institute of Stomatology, Nanjing University from January 1, 2025 to December 15, 2025. Four samples of inflamed dental pulp tissues were taken from patients with pulpitis, and the other four samples of normal dental pulp tissues were collected from healthy third molars that were extracted due to orthodontic or impacted reasons. This study has received informed consent from the patients and has been approved by the Medical Ethics Committee of the Affiliated Stomatological Hospital of Nanjing University Medical School (Number: NJSH-2021NL-006). RNA concentration was detected using the NanoPhotometer N50, and reverse transcription was performed using a cDNA synthesis kit, and primers were synthesized by a biological company (Table 1). The RT-qPCR experiments were performed with GAPDH as the internal reference gene, and the expression level of the key genes was calculated using the 2^−ΔΔCt^ method. ‘GraphPad Prism’ (v 10.1.2)[26] was employed to plot and calculate the P value. Differences between PCR experimental groups were obtained through a t-test (P < 0.05).

**Table 1.**
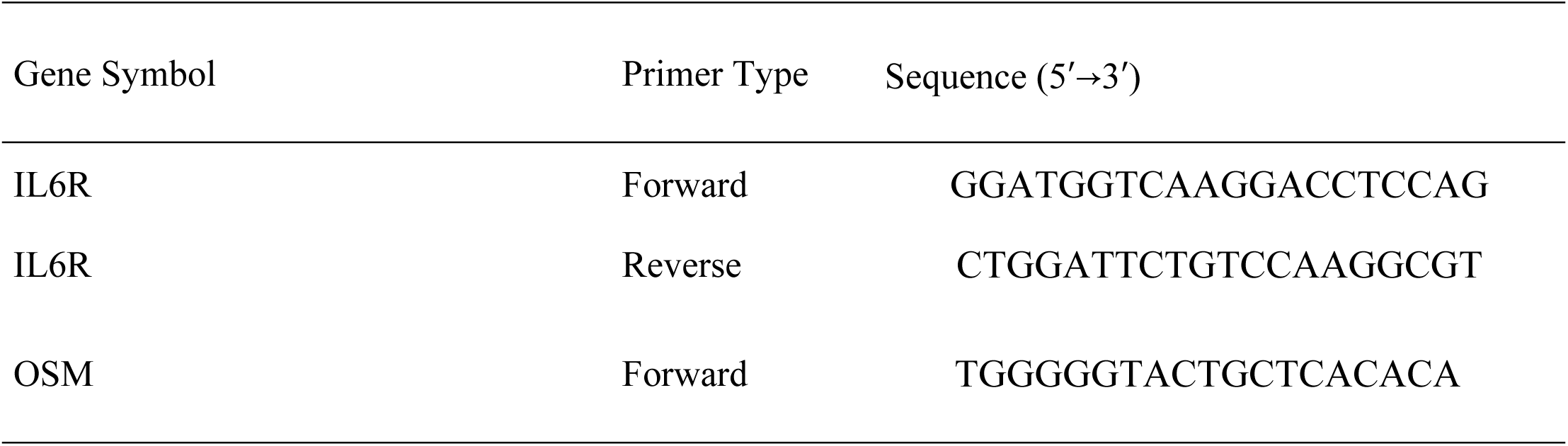
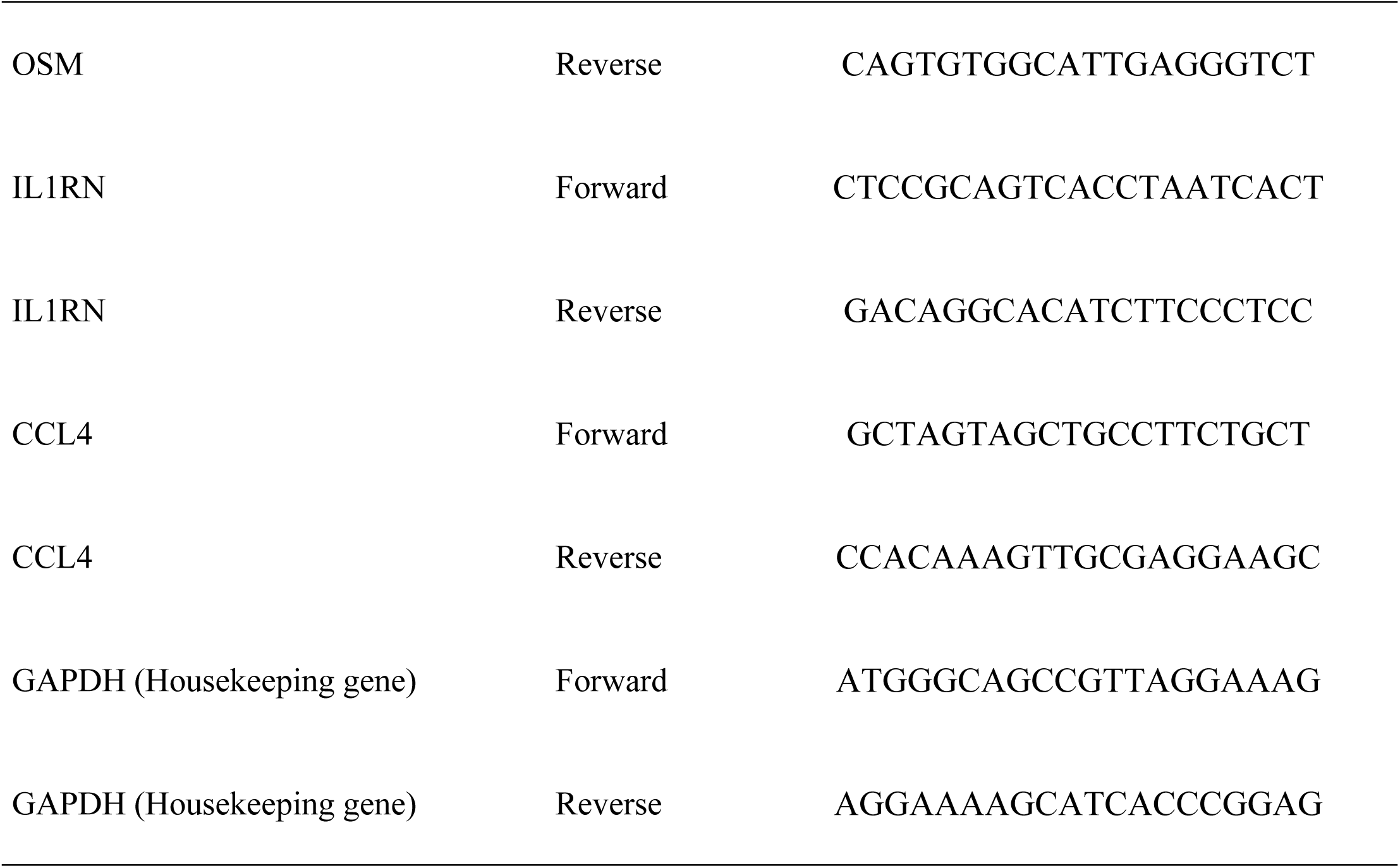
Primer sequences used for RT-qPCR.

### 2.10 Statistical evaluation

Bioinformatics analyses relied on the R programming language (v 4.2.2). Intergroup differences were assessed through the Wilcoxon test, with statistical significance set at P < 0.05.

## 3. Results

### 3.1 There were 10 candidate key genes associated with neuroendocrine regulation

Between pulpitis and control samples in the training set, 1,014 differentially expressed genes were found, with 766 being upregulated and 248 downregulated (**Fig 2A-B**). In order to further identify neuroendocrine regulation-associated DEGs, 1,014 DEGs and 1,173 NER-RGs were subjected to intersection analysis, with 104 intersection genes acquired as candidate genes (**Fig 2C**). Furthermore, GO enrichment analysis showed that (**Fig 2D, S2 Table**). Among them, the biological processes included leukocyte migration and cell chemotaxis; cellular component included membrane microdomain and specific granule; molecular function included cytokine activity, receptor ligand activity, cytokine receptor binding, cytokine binding, chemokine activity (P < 0.05). Moreover, it was shown that the top 5 pathways for KEGG enrichment were viral protein interaction with cytokine and cytokine receptor, TNF signaling pathway, cytokine-cytokine receptor interaction, rheumatoid arthritis, lipid, and atherosclerosis (P < 0.05) (**Fig 2D, S3 Table**). In the PPI network, there were 93 interacting proteins (**Fig 2E**). The top 10 proteins screened by the DMNC algorithm included CCL4, CXCR2, CXCR3, IL6R, etc, and were used as candidate key genes. Among them, IL6R interacted with other proteins most frequently (**Fig 2F**). Overall, the comprehensive analysis offered a wealth of information that could guide further research efforts aimed at understanding the underlying mechanisms of pulpitis and developing innovative therapeutic strategies.

**Fig 2.**
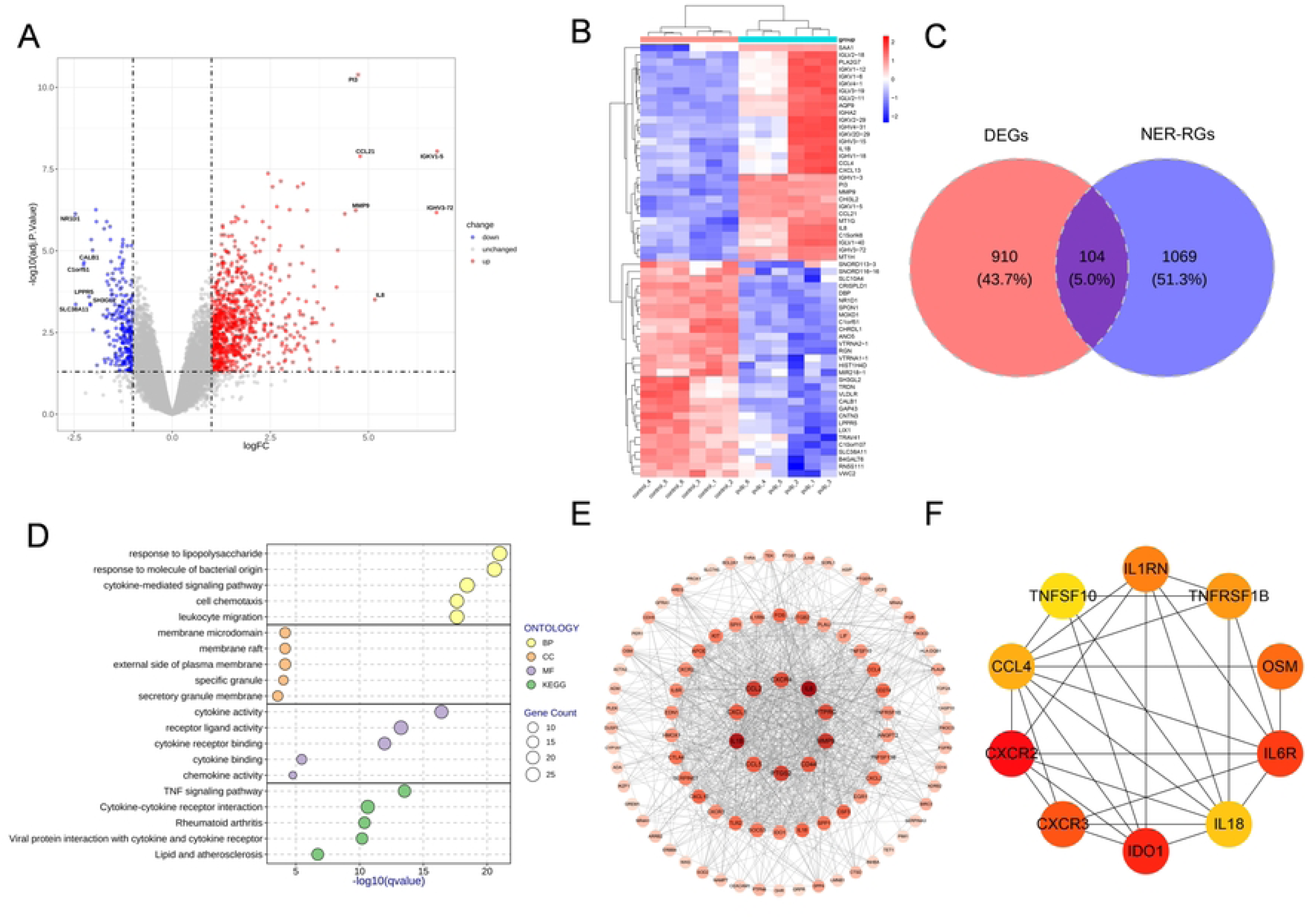
A Volcano plot of differentially expressed genes. B. Heatmap of differentially expressed genes. C. Venn diagram of candidate genes. D. GO and KEGG enrichment analysis. E. Candidate key gene: Protein-Protein Interaction Network. F. The top 10 candidate key gene subnetworks of the DMNC score.

### 3.2 A total of 4 key genes affecting pulpitis were uncovered

In the training set, these 10 candidate key genes showed significant differences in expression between pulpitis and control samples (P < 0.05) (**S4 Table**); in the validation set, only the IL6R, OSM, IL1RN, and CCL4 genes were significantly different between the pulpitis and control samples, and their expression trend was consistent with the training set (P < 0.05), two genes were not expressed in the validation set (**Fig 3, S5 Table**). Therefore, IL6R, OSM, IL1RN, and CCL4 were utilized as key genes.

**Fig 3.**
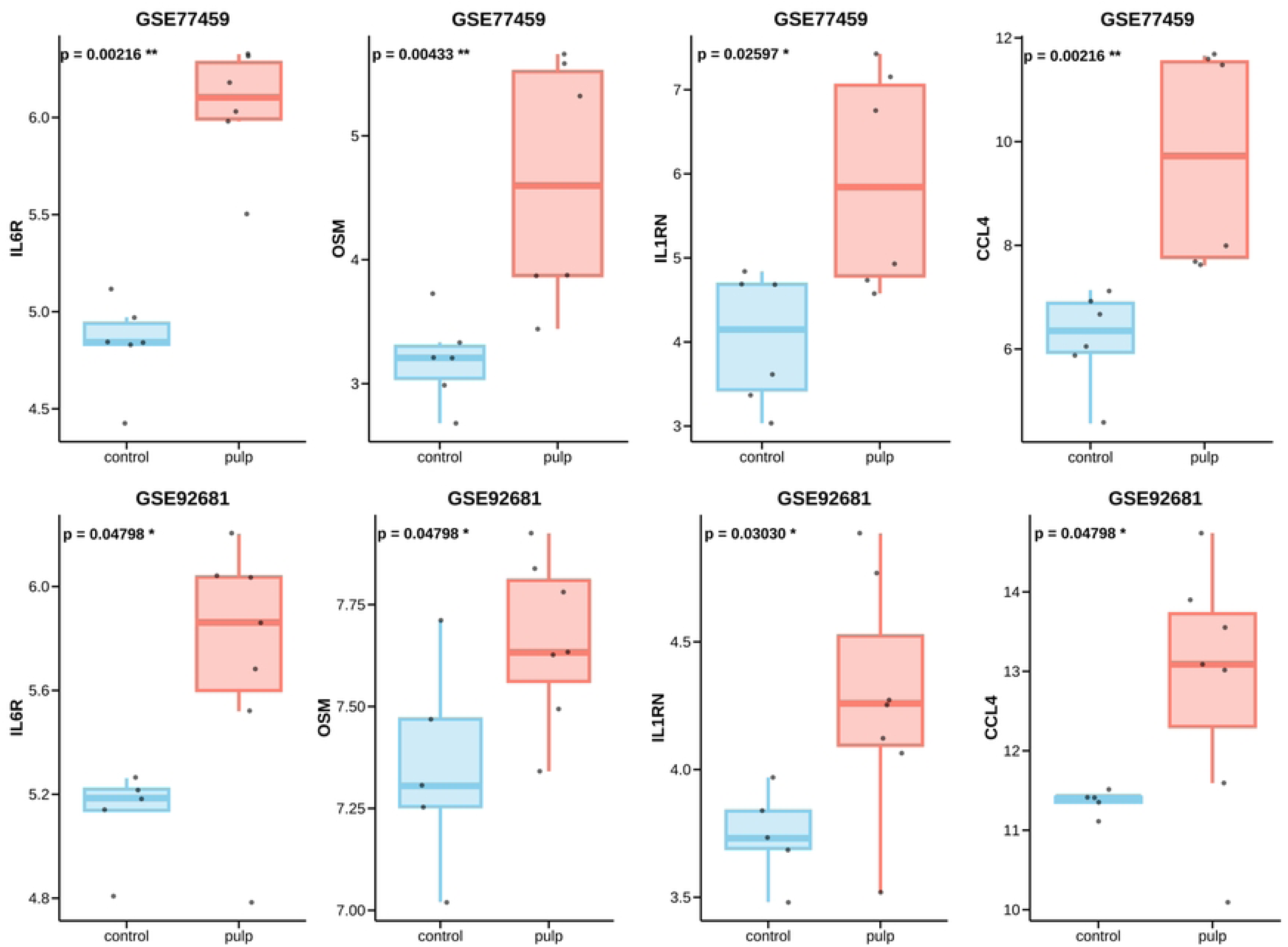
Box plots of expression levels of key genes in the test set and training set.

### 3.3 The regulation of pulpitis was mediated by key genes via multiple pathways

GSEA results for the 4 key genes showed that IL1RN was enriched in 53 pathways, and the top 5 pathways were cytokine-cytokine receptor interaction, spliceosome, Huntington’s disease, olfactory transduction, and chemokine signaling pathway (**Fig 4A, S6 Table**); OSM was enriched in 62 pathways, and the top 5 pathways were hematopoietic cell lineage, oxidative phosphorylation, spliceosome, Huntingtons disease, and cytokine-cytokine receptor interaction (**Fig 4B, S7 Table**); IL6R was enriched in 63 pathways, and the top 5 pathways were oxidative phosphorylation, spliceosome, Huntingtons disease, cytokine-cytokine receptor interaction, and olfactory transduction (**Fig 4C, S8 Table**); CCL4 was enriched in 55 pathways, and the top 5 pathways were Huntingtons disease, cytokine-cytokine receptor interaction, chemokine signaling pathway, olfactory transduction, and oxidative phosphorylation (**Fig 4D, S9 Table**). The common pathways of these 4 key genes were Huntington’s disease and cytokine-cytokine receptor interaction. The above results indicated that the key genes might affect the progression of pulpitis by regulating the occurrence of these pathways.

**Fig 4.**
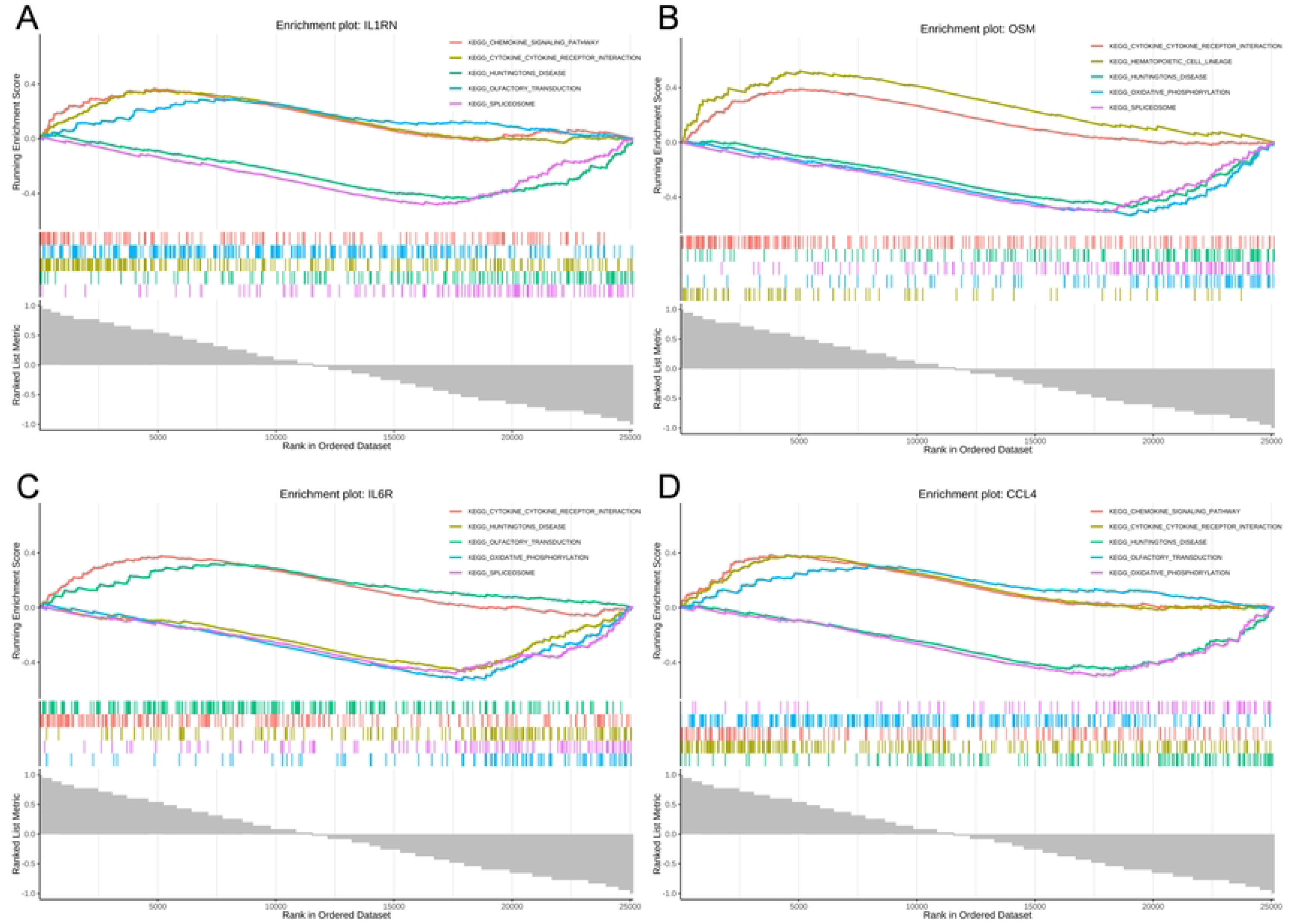
GSEA enrichment analysis of key genes. A. GSEA enrichment analysis of IL1RN. B. GSEA enrichment analysis of OSM. C. GSEA enrichment analysis of IL6R. D. GSEA enrichment analysis of CCL4.

### 3.4 Multiple molecules and drugs might be related to the key genes in pulpitis

A total of 5 RBPs interacting with OSM were identified, namely EIF3A, EIF3D, EIF3G, HNRNPA1, and RBM4. For IL1RN, 15 interacting RBPs were found, including AKAP1, AQR, FAM120A, HNRNPC, and HNRNPK. As for IL6R, 47 interacting RBPs were detected, such as ALYREF, CHTOP, CSTF2, CSTF2T, and ELAVL1. However, no RBP interacting with CCL4 was predicted (**Fig 5A**).

**Fig 5.**
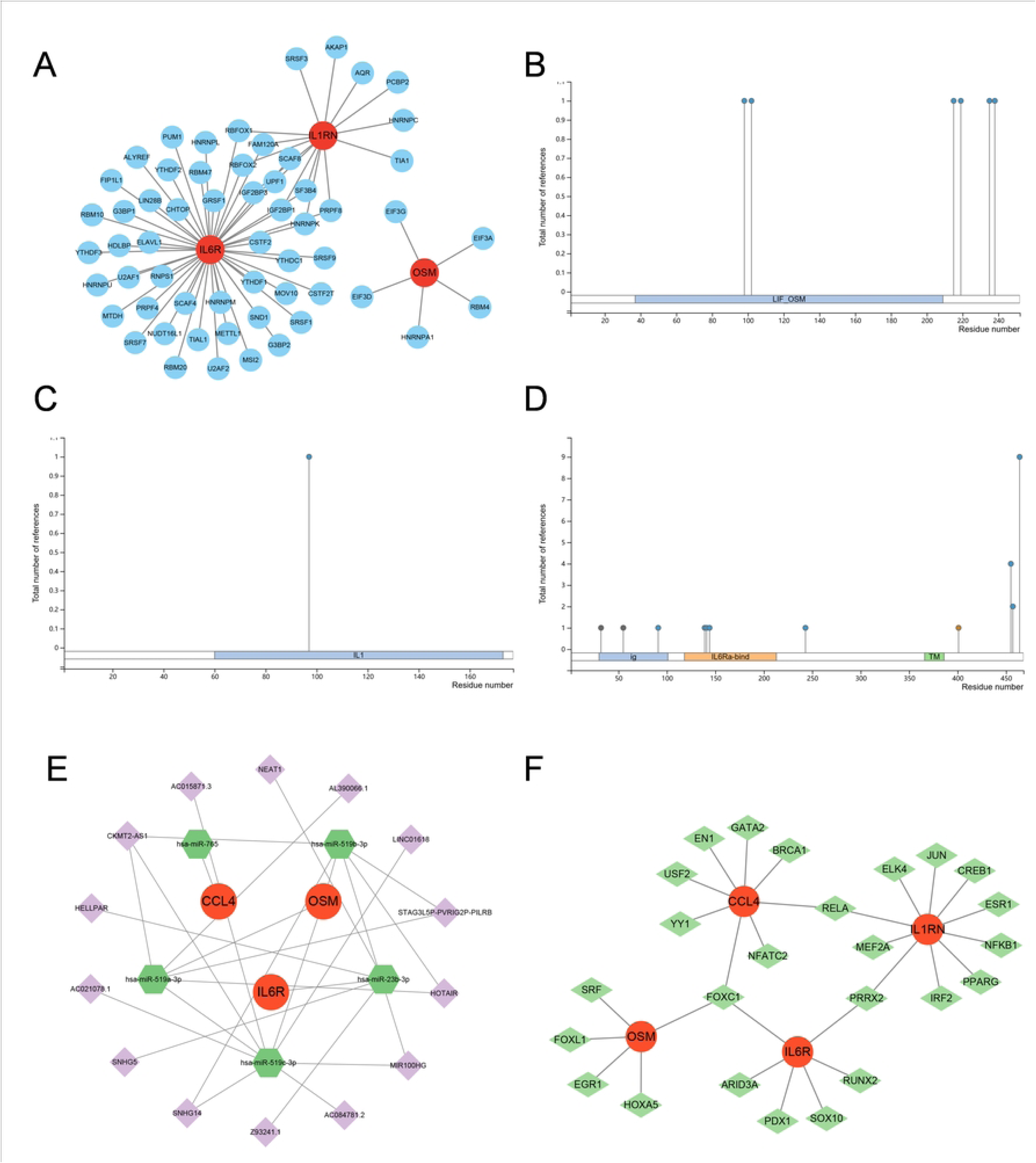
Analysis of RBP regulatory networks and PTM analysis of key genes. A. The RBP regulatory network of key genes. B-D. Post-translational modifications of OSM, IL1RN, and IL6R protein. E. LncRNA-miRNA-mRNA regulatory network. F. mRNA-TFs regulatory network.

The findings regarding the post-translational modifications of the proteins encoded by the 4 key genes indicated that OSM harbored 6 phosphorylation sites (**Fig 5B**). IL1RN had 1 phosphorylation site (**Fig 5C**). IL6R was found to have 8 phosphorylation sites and 1 ubiquitination site, while CCL4 lacked any post-translational modification sites (**Fig 5D**). These results not only facilitated a more in-depth comprehension of the post-translational modification profiles of the key gene-derived proteins but also offered valuable references for the treatment of pulpitis.

The miRNA prediction analysis for the key genes revealed that the microcosm predicted 107 miRNAs, while the Diana Microt database predicted 95 miRNAs. Notably, a total of 5 mRNA-miRNA intersection pairs were identified among the predicted results. The lncRNA-miRNA-mRNA results showed that 3 miRNAs regulated OSM expression, such as hsa-miR-519b-3p, hsa-miR-519a-3p, and hsa-miR-519c-3p, and there were 16 lncRNAs upstream of the miRNAs; hsa-miR-765 regulated CCL4 expression, but no lncRNA was retrieved upstream of the miRNA; hsa-miR-23b-3p regulated IL6R expression, and there were 4 lncRNAs upstream of the miRNA (**Fig 5E**). The results of the TF-mRNA showed that a total of 25 TFs regulated the expression of key genes. Among them, FOXC1 was found to regulate the expression of OSM, IL6R, and CCL4. Meanwhile, PRRX2 was shown to modulate the expression of IL6R and IL1RN. Additionally, RELA was demonstrated to control the expression of CCL4 and IL1RN (**Fig 5F**).

Utilizing the DGIdb database, a total of 19 drugs associated with key genes were identified (**Fig 6**). Regarding IL6R, as many as 12 drugs were predicted, such as sarilumab, satralizumab, and levilimab. For IL1RN, 4 drugs were predicted, including haloperidol decanoate, methotrexate, diacetylrhein, heparin. In the case of CCL4, 3 drugs were predicted, namely cyclosporine, clodronic acid, and epoetin alfa. However, when it came to OSM, no associated drug was retrieved through the database search. Furthermore, drawing on the CTD database, the outcomes of the disease-related scores of the key genes that exceeded 150 were presented (**Fig 6**). Specifically, 4 diseases were linked to OSM, among which were inflammation and weight loss. For IL6R, 6 associated diseases were identified, including weight loss and inflammation. When it came to IL1RN, a total of 33 diseases were correlated with it, such as inflammation and chemical and drug-induced liver injury. In the case of CCL4, 14 related diseases were discovered, like inflammation and necrosis. All 4 key genes were associated with inflammation.

**Fig 6.**
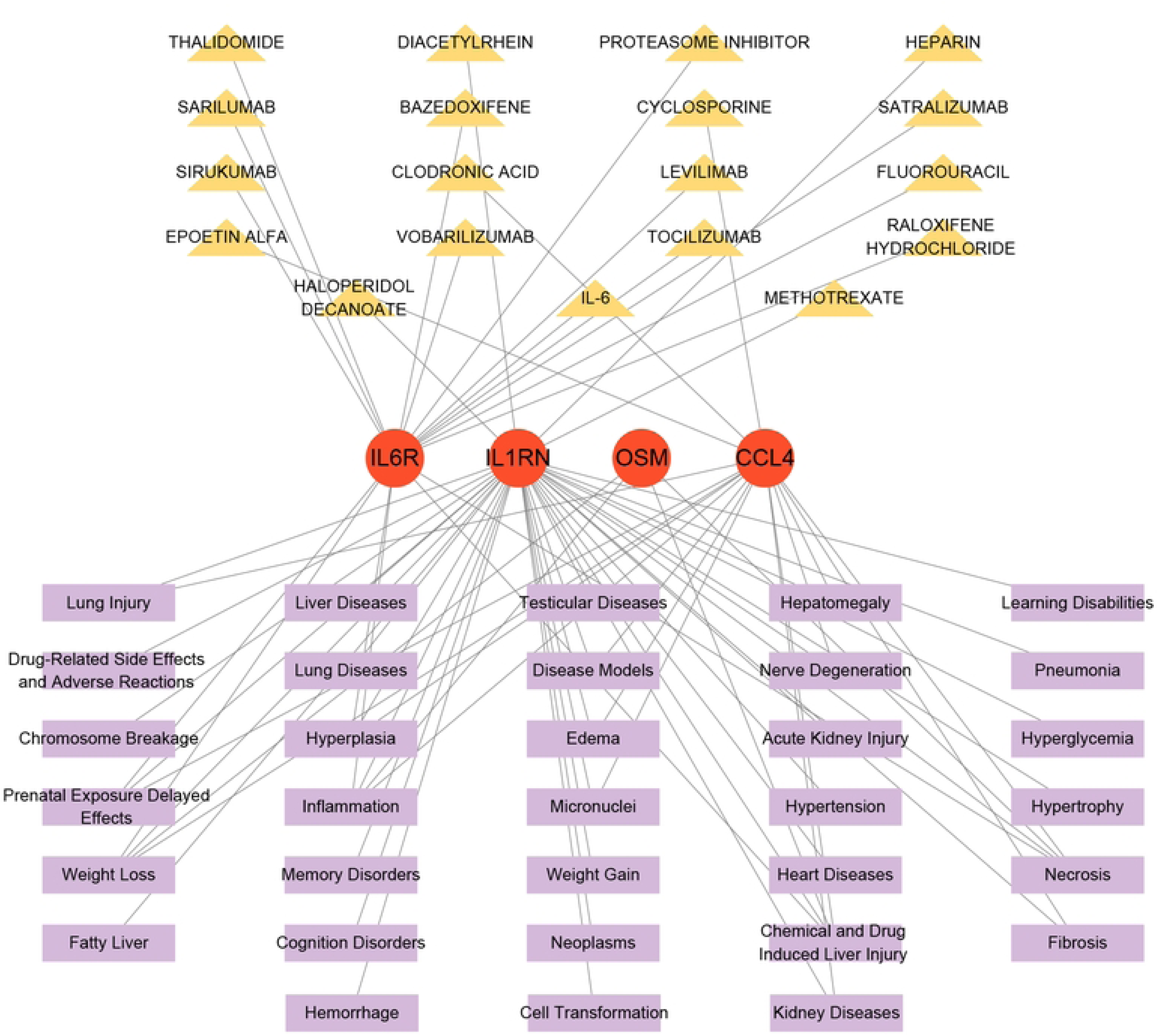
Disease-key gene-drug interaction network.

### 3.5 The expression of key genes in pulpitis and control samples was analyzed

The expression of 4 key genes was detected in clinical samples. The results showed that OSM, IL1RN, and CCL4 were still prominently overexpressed in pulpitis samples compared with control samples (P < 0.05) (**Fig 7**). Although there was no significant difference in IL6R expression, the trend showed higher expression in the pulpitis group. This was in line with the results derived from bioinformatics analysis, verifying the reliability of the bioinformatic analyses.

**Fig 7.**
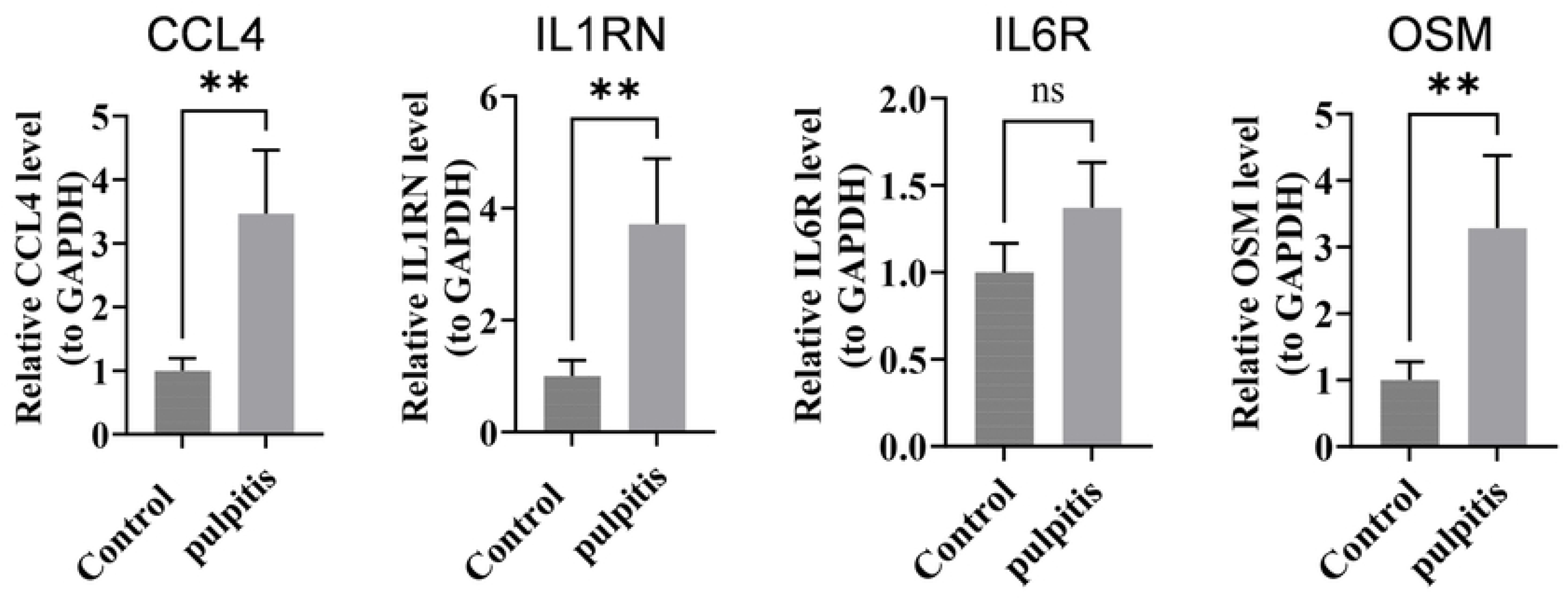
The results of RT-qPCR experiments.

## 4. Discussion

Pulpitis is an inflammatory disease of the dental pulp that arises from the intricate interplay among microbial invasion, immune responses, and neural signaling[2]. During pulpitis development, pulp nerves serve a dual function: they not only mediate pain sensation but also actively promote inflammatory responses through neuropeptide secretion.[27] Moreover, neuroendocrine regulation (NER) is recognized as a critical mechanism in modulating inflammatory pathways and maintaining immune homeostasis25. Given the rich innervation of dental pulp tissue, dysregulation of NER may be intimately linked to pulpitis progression.[28] This study employed transcriptome analysis integrated with RBP regulatory networks, molecular regulatory networks, drug target prediction, and other multi-omics approaches to elucidate the mechanisms underlying the involvement of key genes related to neuroendocrine regulation in pulpitis. These findings enhance our understanding of the pathogenesis of pulpitis and offer novel insights into its therapeutic management.

To further understand the molecular mechanisms, functional enrichment analysis of neuroendocrine regulation-related genes in pulpitis revealed several key pathways underlying disease pathogenesis. Inflammation-related biological processes such as “response to lipopolysaccharide”, “response to molecule of bacterial origin”, and “leukocyte migration” were uncovered via GO analysis.[29] These findings highlight the crucial role of bacterial infection-triggered inflammation in pulpitis development and underscore the importance of immune cell recruitment and activation in inflamed pulp tissue.[30, 31] These GO findings align with KEGG pathway analysis, which revealed enrichment in “TNF signaling pathway” and “cytokine-cytokine receptor interaction” pathways. These pathways can amplify immune cell recruitment and promote sensory neuron sensitization, thereby exacerbating pain perception and tissue damage.[32] The appearance of “ secretory granule membrane” as a cellular component in the functional enrichment suggests that neuroendocrine cells may directly release regulatory molecules into the pulp microenvironment.[33] Collectively, these findings suggest that pulpitis involves diverse biological processes and regulatory pathways that are modulated by neuroendocrine factors. This integrated perspective provides a valuable molecular framework for understanding the pathogenesis of pulpitis and offers an entry point for therapeutic strategies targeting the neuroimmune axis.

This study identified IL6R, OSM, IL1RN, and CCL4 as key genes significantly upregulated in pulpitis through transcriptome analysis, revealing their dual roles in driving inflammatory responses and mediating neuroendocrine regulation. In pulpitis, the downregulation of microRNA-30b can relieve its inhibitory effect on IL6R, leading to high expression of IL6R. After IL-6R binds to IL-6, it can promote the progression of inflammation by activating signaling pathways such as JAK-STAT and MAPK.[34] OSM is upregulated in pulpitis and can act on odontoblasts, fibroblasts, inflammatory cells and endothelial cells, activating related signaling pathways and promoting the release of inflammatory factors and matrix metalloproteinases (MMPs), thereby exacerbating pulp inflammation and tissue damage.[35] The IL1RN gene encodes interleukin-1 receptor antagonist (IL-1ra), which functions as a natural inhibitor of IL-1 signaling. IL-1ra expression is elevated in inflamed pulp tissue, and it competitively binds to IL-1 receptors to block IL-1 β ‘ s pro-inflammatory effects, a key mediator in pulpitis pathogenesis, and helping to modulate the inflammatory respons.[36] CCL4 is a potent chemokine that demonstrates significant upregulation in our pulpitis analysis. While specific mechanistic studies in pulpitis are limited, extensive research in other inflammatory conditions has established CCL4’s role in recruiting monocytes, macrophages, and T lymphocytes to inflammatory sites, thereby amplifying local immune responses and potentially contributing to tissue damage.[2, 37] More importantly, these genes can maintain neuroendocrine homeostasis in physiological stress and inflammatory diseases by directly acting on neuroendocrine organs (such as the hypothalamus and pituitary gland) or indirectly regulating the inflammatory network.[38–41] The consistent up-regulation of these genes makes them potential candidate diagnostic biomarkers. Detecting the elevated mRNA or protein levels of these genes in easily accessible samples such as saliva may provide an effective means for the early non-invasive diagnosis of pulpitis. From a therapeutic perspective, targeted regulation of these genes holds significant intervention potential. Future research should further validate the functional significance and translational application value of these potential targets in pulpitis-specific neuroendocrine models.

To further elucidate the functional mechanisms of these key genes, GSEA was performed and revealed significant enrichment in several important signaling pathways, including cytokine-cytokine receptor interaction, Huntington’s disease pathway, chemokine signaling pathway, and oxidative phosphorylation. These pathways not only play a core regulatory role in the inflammatory response but are also closely related to neuroendocrine regulation (NER), further supporting the potential mechanism of NER in the progression of pulpitis. Prior research has shown that cytokines and chemokines are key to the onset, development, and tissue injury of pulpitis through mediating immune responses, regulating cell migration, and engaging in epigenetic regulatory networks.[42] Additionally, the oxidative phosphorylation pathway, which is central to cellular energy metabolism, influences the balance between inflammatory responses and tissue repair through its effects on immune cell metabolic reprogramming and dental pulp stem cell (DPSC) bioenergetics.[43] The enrichment of Huntington’s disease pathway, which encompasses neurodegeneration, oxidative stress responses, mitochondrial dysfunction, and neuroinflammation mechanisms41, may reflect shared molecular processes between neurodegenerative conditions and pulpal inflammation.[44] The significant enrichment of this pathway suggests that neuroendocrine signaling disorders in pulp neurons may exacerbate neurogenic inflammation. Moreover, the GSEA results are highly consistent with the GO and KEGG analysis results, further validating these genes as core factors in the neuroendocrine regulatory network related to pulpitis. Therefore, targeting these key genes or their related pathways may help alleviate neuroendocrine disorders in pulpitis and provide new therapeutic strategies for maintaining pulp vitality.

To elucidate the regulatory mechanisms underlying key gene functions and their molecular network interactions, we performed RNA-binding protein (RBP) network analysis, protein post-translational modification (PTM) analysis, and molecular regulatory network constructionPrevious studies have shown that RNA-binding proteins (RBPs) play a crucial role in the occurrence, development, and repair of pulpitis.[45] Based on RBP regulatory network analysis of the key genes, this study identified multiple RBPs that participate in pulpitis progression and may have potential implications for stress-related cellular responses. Among them, ELAVL1 and HNRNPA1 can maintain the stability of inflammatory factor mRNA by binding to specific regions.[46, 47] RBM4 and HNRNPC can affect the generation of inflammatory-related protein subtypes by regulating the splicing of precursor mRNA.[48, 49] EIF3A and EIF3D can participate in the assembly of translation complexes, thereby regulating the translation efficiency of inflammatory factors.[50, 51] HNRNPK and CSTF2 can indirectly regulate the activity of inflammatory signaling pathways by interacting with lncRNA or miRNA.[52, 53] Additionally, CHTOP and AKAP1 can indirectly affect the homeostasis of the neuroendocrine system by regulating inflammatory responses, metabolic processes, and cellular stress states.[54, 55] Notably, HNRNPA1, HNRNPC, HNRNPK, and ELAVL1 can also influence the occurrence, differentiation, and malignant transformation of various neuroendocrine tumors by regulating gene expression stability, signaling pathway activity, and the immune microenvironment, further revealing the core functions of key gene-related RBPs in neuroendocrine regulation.[56–59]

Furthermore, our research has found that phosphorylation is the most significant form of protein post-translational modification (PTM) among these key genes. On the one hand, phosphorylation modification can drive the development of pulp inflammation by regulating the core molecules Inhibitor of kappa B alpha (IκBα) and p65 in the Nuclear Factor kappa B (NF-κ B) signaling pathway.[60] On the other hand, phosphorylation modification can also provide an important molecular basis for the homeostatic regulation of the neuroendocrine system by modulating the conformation and function of key proteins such as AnxA2.[61]

To comprehensively understand gene regulation, molecular regulatory network analysis was performed to elucidate the interactions among key genes in pulpitis. Among them, the core miRNAs - miR-519, miR-765, and miR-23 - are important regulatory factors of the inflammatory response.[62–64] Notably, miR-519 family members target OSM expression, with 16 lncRNAs potentially acting as competing endogenous RNAs (ceRNAs) to modulate this regulatory interaction. In contrast, the regulation of CCL4 by miR-765 does not involve long non-coding RNAs, indicating that this chemokine has a rapid and direct post-transcriptional regulatory pattern in pulpitis. From the perspective of transcription factors, this study found that transcription factors FOXC1, RELA, and PRRX2 all can cooperatively regulate multiple targets. FOXC1 is involved in the regulation of vascular development and neuronal injury,[65, 66] suggesting its possible association with microvascular dysfunction and neurogenic inflammation in the dental pulp. RELA is involved in the regulation of inflammatory responses and neuropathic pain,[67] suggesting that it may mediate local inflammation in the dental pulp and participate in the sensitization process of pulp neuralgia. PRRX2 is involved in the differentiation of mesenchymal cells,[68] and the dental pulp tissue contains a large number of cells derived from mesenchyme,[69] indicating that it may participate in the occurrence and development of pulpitis by influencing the differentiation process of pulp cells. Collectively, these transcription factors may coordinate inflammatory gene expression programs in pulpitis through their multi-target regulatory networks, potentially contributing to disease progressionThis study analyzes the functional regulatory mechanisms of these key genes from multiple levels, which is of great significance for the in-depth exploration of the pathogenesis of pulpitis. Through targeted intervention measures such as RBP, phosphorylation modification, and specific miRNA, it is expected that new treatment strategies can be developed in future clinical treatment, thereby improving the prognosis of patients with pulpitis.

Drug prediction and disease association analyses were performed to identify compounds targeting the key gene products and their clinical relevance. The results showed that the number of predicted drugs for the IL6R gene was the largest. Among them, anti-IL6R monoclonal antibodies represented by sarilumab and satralizumab can effectively alleviate inflammatory responses, pain sensitization, and nerve damage by inhibiting the IL-6 signaling pathway.[70–73] For IL1RN, the predicted drug methotrexate has the effect of inhibiting the proliferation of inflammatory cells and the formation of new blood vessels, which may help reduce the infiltration of inflammatory cells in the pulp tissue and alleviate vascular dilation and tissue damage.[74, 75] Diacetylrhein may create a more favorable microenvironment for pulp tissue repair by inhibiting the activity of NLRP3 inflammasome and reducing alveolar bone resorption.[76, 77] Haloperidol decanoate has the potential to block pain signal transmission.[78] However, these drugs have certain risks of adverse reactions such as bone marrow suppression, liver toxicity, and extrapyramidal reactions, which limit their safety for systemic application. Given these limitations, targeted local delivery approaches might theoretically offer safer alternatives, though such applications would require extensive preclinical and clinical validation. Disease association analysis further indicated that all four key genes were closely related to inflammation. Among them, the extensive disease association of IL1RN suggests that it may have a systemic regulatory role; the significant association of CCL4 with “necrosis” is consistent with the pathological characteristics of irreversible pulp damage, while the association of OSM and IL6R with “weight loss” suggests that pulpitis may involve neuroendocrine metabolic disorders.

Finally, the RT-qPCR experiment verified that there were statistically significant abnormal expressions of three key genes related to neuroendocrine regulation in the pulpitis samples; although the expression level of IL6R did not reach a significant difference, it showed an upward trend in the pulpitis group. This result was consistent with the transcriptome sequencing analysis, further supporting the reliability of the research results of this study.

Despite these findings, it should be acknowledged that the current study has certain limitations. The overall outcomes are dependent on data quality, algorithm accuracy, and analytical assumptions. There may be problems such as data noise, poor interpretability due to algorithm complexity, and false-positive and false-negative results. Going forward, further exploration of gene functions can be conducted at the multi-omics level, such as studying the expression of key genes in different pulpitis cell subtypes at the single-cell level, or exploring the epigenetic changes and regulatory mechanisms of these key genes.

## 4. Conclusion

This study, through transcriptome analysis and experimental verification, identified four key genes related to neuroendocrine regulation in pulpitis and investigated their mechanism of action in pulpitis, providing a new theoretical basis and reference value for the treatment of pulpitis.

## Acknowledgments

No Acknowledgments

## Supporting information

S1 Table. 1,173 neuroendocrine regulation-related genes.

S2 Table. GO enrichment analysis.

S3 Table. KEGG enrichment analysis.

S4 Table. The expression difference of candidate genes in the training set.

S5 Table. The expression difference of candidate genes in the validation set.

S6-S9 Table. GSEA analysis of 4 key genes.

## Author contribution

**Conceptualization:** Huimin Jin, Yawei Wang, Ting Guo.

**Data curation:** Huimin Jin, Yawei Wang.

**Formal analysis:** Huimin Jin, Yawei Wang.

**Investigation:** Huimin Jin, Yawei Wang.

**Methodology:** Huimin Jin, Yawei Wang.

**Resources:** Huimin Jin, Aoteng Sun, Yun Liu.

**Software:** Huimin Jin, Yawei Wang.

**Visualization:** Huimin Jin, Yawei Wang.

**Writing – original draft:** Huimin Jin, Yawei Wang, Aoteng Sun, Yun Liu.

**Supervision:** Aoteng Sun, Ting Guo.

**Validation:** Aoteng Sun, Yun Liu.

**Project administration:** Ting Guo.

**Writing – review & editing:**Ting Guo.

## Data availability statement

The relevant datasets (GSE77459,GSE92681) utilized in this study were publicly available and obtained from Gene Expression Omnibus (GEO, https://www.ncbi.nlm.nih.gov/geo/). The raw da ta, R code, and RT-qPCR experimental data related to this study are available in the Zenodo repository, accessible via DOI: https://doi.org/10.5281/zenodo.19563399.

## Ethics approval statement

This study has received informed consent from the patients and has been approved by the Medical Ethics Committee of the Affiliated Stomatological Hospital of Nanjing University Medical School (Number: NJSH-2021NL-006).

